# Ischemic Stroke in COVID-19: An Urgent Need for Early Identification and Management

**DOI:** 10.1101/2020.05.25.20111047

**Authors:** Dinesh V. Jillella, Nicholas J. Janocko, Fadi Nahab, Karima Benameur, James G. Greene, Wendy L. Wright, Mahmoud Obideen, Srikant Rangaraju

## Abstract

**Objective:** In the setting of the Coronavirus Disease 2019 (COVID-19) global pandemic caused by SARS-CoV-2, a potential association of this disease with stroke has been suggested. We aimed to describe the characteristics of patients who were admitted with COVID-19 and had an acute ischemic stroke (AIS).

**Methods:** This is a case series of PCR-confirmed COVID-19 patients with ischemic stroke admitted to an academic health system in metropolitan Atlanta (USA) between March 24^th^,2020, and May 5^th^, 2020. Demographic, clinical, and radiographic characteristics were described.

**Results:** Of 124 ischemic stroke patients admitted during this study period, 8 (6.5%) were also diagnosed with COVID-19. The mean age of patients was 64.3 ± 6.5 years, 5 (62.5%) male, mean time from last-normal was 4.8 days [SD 4.8], and none received acute reperfusion therapy. All 8 patients had at least one stroke-associated co-morbidity. The predominant pattern of ischemic stroke was embolic; 3 were explained by atrial fibrillation while 5 (62.5%) were cryptogenic. In contrast, cryptogenic strokes were seen in 20 (16.1%) of 124 total stroke admissions during this time.

**Conclusions:** In our case series, ischemic stroke affected COVID-19 patients with traditional stroke risk factors with an age of stroke presentation typically seen in non-COVID populations. We observed a predominantly embolic pattern of stroke with a higher than expected rate of cryptogenic strokes and with a prolonged median time to presentation and symptom recognition limiting the use of acute reperfusion treatments. These results highlight the need for increased community awareness, early identification, and management of AIS in COVID-19 patients.

## Introduction

The United States has seen an exponential rise in COVID-19 infection cases recently, resulting in over 1.7 million cases and > 100,000 deaths. In the setting of the COVID-19 pandemic, there have been reports of its association with various neurological disorders including stroke (1, 2). Stroke has been reported in 5.7 % of patients with severe COVID-19 infections and 0.8 % of patients with non-severe infection in an Asian study (1). Previous studies have reported the role of infectious agents in stroke pathogenesis (3). Based on the underlying hypercoagulability and increased incidence of thrombotic events in severe COVID-19 patients, increased risk of stroke, particularly of embolic etiology, may be predicted. A recent case series of 5 young COVID-19 patients with embolic strokes with few/no traditional risk factors supports this idea (4). However, there are very limited data on the characteristics and frequency of stroke in COVID-19. In this case series from a large academic hospital system in the Southern United States, we report clinical and radiographic characteristics of eight acute ischemic stroke (AIS) patients with COVID-19 infection.

## Methods

### Study Design

This is a retrospective study of adult patients aged 18 years or above admitted to Emory Healthcare hospitals in Atlanta, Georgia, USA with a diagnosis of COVID-19 and identified to have an acute ischemic stroke at initial presentation or during hospital admission. The Institutional Review Board of Emory University approved this retrospective study.

### Inclusion and Exclusion Criteria

Adult patients aged 18 years or above at the time of PCR-confirmed COVID-19 admission from March 24^th^, 2020 to May 5^th^, 2020 were included. Diagnosis of acute ischemic stroke based on clinical and radiological confirmation at presentation or during admission was required for inclusion. Patients were excluded if they did not have either clinical or radiological confirmation on review of the patient records.

### Study Variables

We captured demographic variables including age, sex, and race. Clinical variables collected include vascular risk factors such as diabetes mellitus (DM), hyperlipidemia, hypertension, congestive heart failure (CHF) and atrial fibrillation; deep vein thrombosis (DVT) or pulmonary embolism (PE); ischemic stroke-specific features like etiology as per TOAST guidelines (5); hemispheric laterality and location of stroke; neurological symptomatology; therapeutic interventions used including intravenous tissue plasminogen activator (iv-tPA) or mechanical thrombectomy (MT); stroke outcome measures including modified Rankin Scale (mRS at discharge), admission to the intensive care unit (ICU) and the duration of ICU stay; non-neurological presenting symptoms and diagnoses; brain imaging and laboratory characteristics in COVID-19 infection and therapies used.

## Results

A total of 124 ischemic stroke patients were admitted during this time-period including 8 (6.5%) who were diagnosed with COVID-19. Mean age was 64 +/-5.7 years, 5 (62.5%) were male and (62.5%) were Black Americans. History of DM was present in 6 (75%) patients and hypertension was present in 7 (88%) patients and all patients had at least one medical comorbidity (DM, hypertension, atrial fibrillation, or hyperlipidemia). The predominant pattern of ischemic stroke observed was embolic (8 of 8) and a clear cardio-embolic cause was identified for 3 cases (atrial fibrillation or flutter). Four patients had D-dimer levels of > 3,000 ng/mL with 3 of these having a level > 60,000 ng/mL at admission, and 3 having deep venous thrombosis (DVT) or pulmonary embolism (PE). In comparison to the high rate of embolic pattern cryptogenic (stroke of undetermined etiology per the TOAST criteria) (5) stroke etiology in this case series, 20 (16.1 %) strokes were deemed cryptogenic out of all the non-COVID-19 stroke admissions across all hospitals of the Emory Healthcare System. Baseline characteristics and clinical and investigational characteristics of these 8 AIS patients with COVID-19 infection are described in Table 1.

**Table 1:** Clinical and radiographic characteristics of AIS patients with COVID-19

|  | **Patient 1** | **Patient 2** | **Patient 3** | **Patient 4** | **Patient 5** | **Patient 6** | **Patient 7** | **Patient 8** |
| --- | --- | --- | --- | --- | --- | --- | --- | --- |
| **Demographics:** |  |  |  |  |  |  |  |  |
| Age | 50s | 60s | 70s | 50s | 60s | 60s | 60s | 60s |
| Sex | Female | Male | Male | Female | Male | Male | Male | Female |
| Race | Black | Unknown | White | Black | Black | White | Black | Black |
| **Stroke Risk Factors/Associated Medical Conditions:** |  |  |  |  |  |  |  |  |
| Diabetes | Yes | No | No | Yes | Yes | Yes | Yes | Yes |
| Hypertension | Yes | No | Yes | Yes | Yes | Yes | Yes | Yes |
| Hyperlipidemia | No | No | Yes | No | No | Yes | Yes | No |
| Atrial Fibrillation or Flutter | No | Yes | No | No | Yes | No | Yes | No |
| DVT/PE | Yes | No | No | No | No | Yes | No | Yes |
| Stroke Characteristics: |  |  |  |  |  |  |  |  |
| Etiology | Cryptogenic | Cardio-embolic | Cryptogenic | Cryptogenic | Cardio-embolic | Cryptogenic | Cardio-embolic | Cryptogenic |
| Location<br>Anterior versus Posterior | Anterior | Anterior and Posterior | Anterior | Anterior | Anterior and Posterior | Anterior | Anterior | Anterior |
| Side | Bilateral | Bilateral | Right | Left | Bilateral | Left | Left | Right |
| Acute Therapy | No tPA or EVT | No tPA or EVT | No tPA or EVT | No tPA or EVT | No tPA or EVT | No tPA or EVT | No tPA or EVT | No tPA or EVT |
| Time from COVID-19 manifestations to stroke symptom onset or identification in days | 11 | 6 | 25 | 0.75 | 7 | 14 | 2 | 2 |
| Last known normal in days | 11 | 6 | 0.5 | 0.75 | 2 | 14 | 2 | 2 |
| Primary Neurological Symptoms | Encephalopathy | Coma | Bilateral Hearing Loss | Aphasia, Right-sided Weakness | Bilateral Lower Extremity Weakness | Obtundation | Aphasia, Encephalopathy | Left-sided weakness and Right Gaze Deviation |
| Cerebral Imaging<br>Figure 1 (A to H) | A) MRI Brain: Small infarctions in the bilateral corona radiata, centrum semiovale, splenium of the corpus callosum, and left temporal lobe involving the bilateral MCA territories. | B) CT Head: Infarct in the left parietal lobe and two infarcts in the frontal and occipital lobes Involving the MCA and PCA territories. | C) MRI Brain: Right temporal lobe infarction of the insular and sub-insular cortex involving the MCA territory. | D) CT Head: Left frontal lobe infarction involving the ACA territory | E) MRI Brain: Small Infarctions involving the bilateral MCA territories. | F) CT Head: Left paramedian occipital lobe and parietal lobe infarction involving the PCA and MCA territories | G) CT Head: Left temporal and parietal lobe infarction involving the MCA territory | H) CT Head: Right frontal, temporal and parietal lobe infarctions involving the MCA territory |
| COVID-19 Characteristics: |  |  |  |  |  |  |  |  |
| Clinical Features: | Hypoxic Respiratory Failure | Respiratory and Cardiogenic Shock | Fatigue | Shortness of Breath, Encephalopathy | Cough, Fever | Hypoxi, Shock, Encephalopathy | Cough, Encephalopathy | Hypoxic Respiratory Failure |
| COVID-19 Therapy: | Hydroxychloroquine | None | None | Hydroxychloroquine | Remdesivir | None | None | None |
| General Therapeutics: |  |  |  |  |  |  |  |  |
| ACE /ARB use | No | No | No | No | Yes | Yes | Yes | No |
| Laboratory Parameters on initial presentation: |  |  |  |  |  |  |  |  |
| WBC (10E3/mcL) | 9.5 | 15.1 | 8.9 | 8.4 | 9.3 | 17.9 | 3.5 | 19.1 |
| Platelet count (10E3/mcL) | 318 | 302 | 255 | 291 | 409 | 337 | 156 | 466 |
| CRP (mg/L) | 182 | 27.5 | 3.3 | 236.4 | 126.1 | 104.2 | 59 | 72.9 |
| Ferritin (ng/ml) | 549 | NA | 433 | NA | NA | NA | 571 | 15 |
| Fibrinogen (mg/dl) | 861 | 571 | NA | NA | 580 | NA | NA | 765 |
| D-Dimer (ng/ml) | 1934 | 4724 | 781 | > 60,000 | 2119 | > 60,000 | 833 | > 60,000 |
| INR | 1.12 | 1.2 | 1.12 | 1.34 | 1.62 | 1.3 | 1.24 | 1.33 |
| LDL (mg/dl) | 46 | UTC | NA | 121 | 22 | 78 | 33 | 55 |
| Triglycerides (mg/dl) | 87 | 301 | NA | 178 | 59 | 94 | 46 | 232 |
| HgA1c (%) | 11.3 | 6.1 | NA | 8.6 | 6.4 | 5.5 | 12.1 | 6.6 |
| Outcomes: |  |  |  |  |  |  |  |  |
| ICU Admission | Yes | Yes | No | Yes | Yes | Yes | No | Yes |
| ICU LOS (days) | 21 | 12 | - | 5 | 1 | 9 | - | 11 |
| mRS on Admission | 0 | 0 | 1 | 0 | 3 | 3 | 1 | 0 |
| mRS at Discharge* | 3 | 6 | 1 | 3 | 4 | 5 | 2 | 6 |

DVT/PE – Deep venous thrombosis/Pulmonary Embolus; ivtPA – intravenous tissue plasminogen activator; MT – Mechanical thrombectomy; MRI – Magnetic Resonance Imaging; CT – Computed Tomography; ACA – Anterior Cerebral Artery; MCA – Middle Cerebral Artery; PCA – Posterior Cerebral Artery; ACE/ARB – Angiotensin-Converting Enzyme/Angiotensin Receptor Blocker; WBC – White blood count; CRP: C-reactive protein, INR- International Normalized Ratio, LDL- Low-density Lipoprotein; UTC = unable to be calculated; MRS – Modified Rankin Scale; ICU – Intensive Care Unit; NA = Not available, LOS – length of stay;

**Figure 1.**
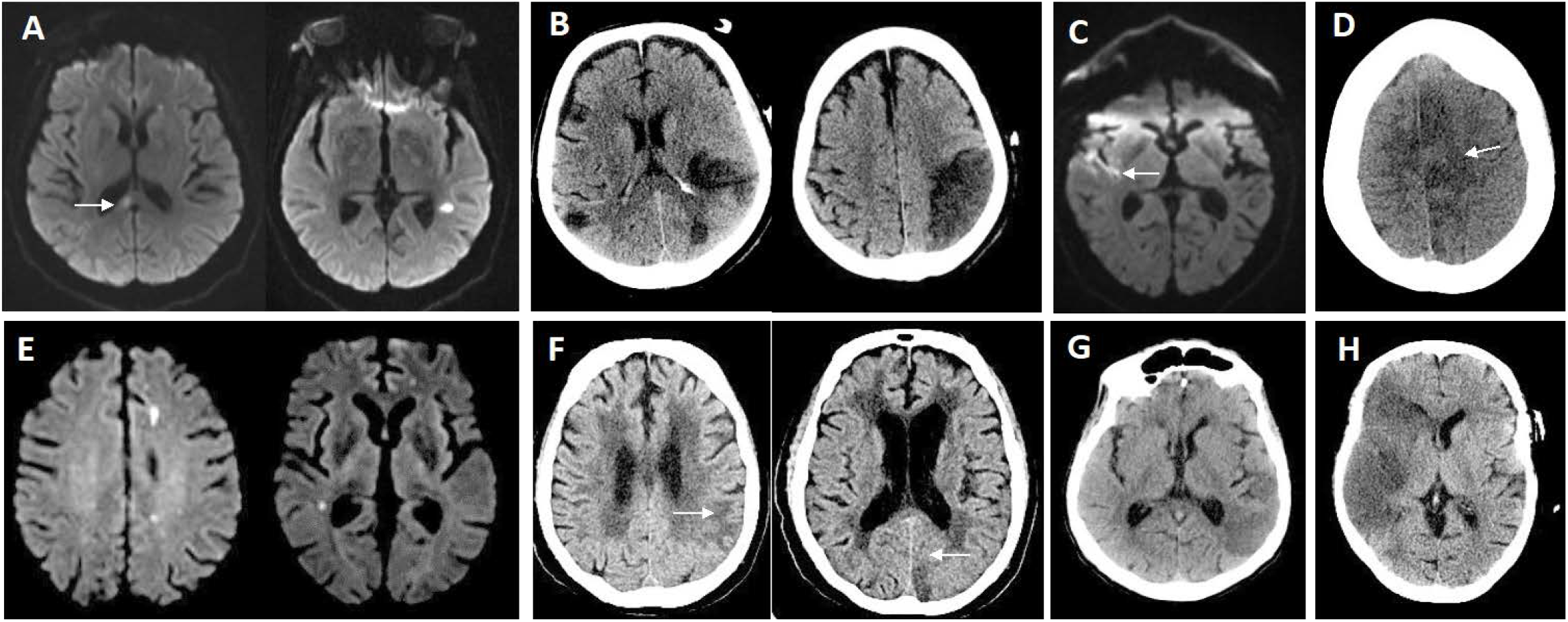
Imaging characteristics of ischemic stroke in COVID-19 patients. 1A: MRI of the brain showing areas of infarction in the right corpus callosum and left temporal lobe on the diffusion-weighted sequence. 1B: CT brain showing hypodensities in the bilateral middle cerebral artery territories. 1C: MRI brain showing a right-sided temporal lobe infarction on the diffusion-weighted sequence. 1D: CT head showing hypodensity in the left anterior cerebral artery territory. 1E: MRI brain showing areas of infarction in the left frontal and right temporal lobes on the diffusion-weighted sequence. 1F: CT head showing hypodensities correlating with infarctions in the left parietal and occipital lobes. 1G: CT head showing a hypodensity in the left temporal lobe 1H: CT head showing a large infarction in the right middle cerebral artery territory.

## Discussion

COVID-19 infection caused by the SARS-CoV2 virus is associated with multi-system dysfunction including neurological involvement, systemic inflammation, and hypercoagulability. Neurological involvement with stroke as a manifestation was reported in 6 (2.8%) patients with COVID-19 based on an Asian study of 214 patients. Ischemic stroke constituted a majority of these patients with 5 of the 6 having a diagnosis of ischemic stroke (1).

Infectious pathogens have been described to contribute to stroke causation via various mechanisms including accelerated atherosclerosis, vasculitis, vasculopathy, and coagulopathy (3). Coagulopathy/hypercoagulable state specifically has been postulated to be one of the important mechanisms of COVID-19’s clinical effects (6, 7). Its possible effect on stroke causation is still unclear although some recent case reports in young patients with few/no comorbid risk factors potentially point towards an association (4). In our study, COVID-19 patients with AIS constituted 6.5% of the ischemic stroke admissions in our healthcare system. All the patients had an embolic pattern stroke noted on imaging and without a clear source of the embolism identified in 5 of the 8 (62.5%) meeting the criteria for cryptogenic strokes that contribute to 10-30% of all stroke cases and lower in centers with advanced testing capabilities (8). Cryptogenic strokes overall constituted 16.1 % at all our stroke centers, considerably lower than that observed in our case series. 3 patients (none of whom had a diagnosis of atrial fibrillation or flutter) also had DVT or PE. Of these 3, 2 patients had D-dimer elevations > 60,000 ng/ml. Based on this information, COVID-19 associated hypercoagulable state stemming from the pro-inflammatory processes (6, 7) could be hypothesized as a possible contributory mechanism for the higher prevalence of embolic cryptogenic stroke presentations in our series. Of note, a recent study from the New York Healthcare System also reported a high prevalence of cryptogenic strokes and suggested a possible association with hypercoagulability (9), although further prospective studies are needed to confirm these findings considering the limitations of retrospective studies.

One patient had a prior diagnosis of atrial fibrillation while two had new diagnoses of atrial fibrillation/flutter during this admission that could be causal in the stroke etiologies in these three patients. Active infectious processes triggering atrial fibrillation events have been described (10) and increasing reports suggest a cardiac involvement with COVID-19 infections (11, 12). Of note, none of the cardiac arrhythmias were observed in patients who received hydroxychloroquine.

All except one patient in our study had a diagnosis of either DM or hypertension (most having both), and the one patient who did not have a prior diagnosis of DM, was found to be pre-diabetic on testing. Although these could be argued as potential confounders from a stroke etiologic standpoint, the presence of these comorbid conditions has been associated with a higher risk of having a COVID-19 infection, and with greater disease severity (13). In this setting, it needs to be ascertained if the presence of hypertension, DM and related upregulation of the ACE2 could contribute to an increased risk of stroke in these patients. In our study, we observed that the average age of COVID-19 stroke presentation is in line with the nation-wide stroke average age of ~ 70.

Acute ischemic stroke therapy has greatly evolved over the last two decades with excellent clinical outcomes reported in recent years with reperfusion strategies of ivtPA and MT. Unfortunately, none of the AIS patients in our series received reperfusion therapy and with primary reasons being either delayed presentation or recognition of atypical stroke symptoms. The diverse constellation of neurological symptoms ranging from encephalopathy to being comatose to having bilateral weakness and atypical bilateral hearing loss rather than the typical findings of cortical involvement or focal motor manifestations along with severe systemic dysfunction compromising the clarity of neurological manifestations can be presumed to be contributory in this delayed stroke recognition. COVID-19 patients with cardio-pulmonary dysfunction are also generally intubated with paralytics and sedatives on board, along with strict isolation measures that could potentially cause delays in stroke recognition. There need to be protocols in place ensuring sedation holidays that can help counter this issue along with more frequent clinical monitoring. Delayed presentations could be a result of patients waiting longer due to self-isolation or quarantine precautions that have led to a general decline in acute stroke evaluations across the country (14). This highlights the need to increase community awareness regarding stroke symptoms and the need to ensure rapid evaluation in the ED to facilitate early stroke treatments and limit disability.

## Limitations

This is a retrospective case series that is limited by the small number of cases and hence, the results are mainly observational and causal association of COVID-19 infection with AIS cannot be ascertained.

## Conclusion

In our case series, ischemic stroke affected COVID-19 patients at an age of stroke presentation typically seen in non-COVID populations. We observed a predominantly embolic pattern of stroke with a higher than expected rate of cryptogenic strokes and with a prolonged median time to presentation/symptom recognition limiting the utilization of acute reperfusion therapy These results highlight the need for community awareness, early identification, and management of AIS in COVID-19 patients. Further studies to determine the effects of COVID-19 associated coagulopathy on ischemic stroke risk as well as the interactions between COVID-19 and other known stroke risk factors are warranted.

## Data Availability

All data owned by authors and de-identified information can be considered for sharing upon request via the correspondent author through appropriate channels.

